# Higher Frequency and Mortality Rate of Antimicrobial-resistant Bloodstream Infections in Tertiary-care Hospitals Compared with Secondary-care Hospitals in Thailand

**DOI:** 10.1101/2023.02.07.23285611

**Authors:** Cherry Lim, Viriya Hantrakun, Preeyarach Klaytong, Chalida Rangsiwutisak, Ratanaporn Tangwangvivat, Chadaporn phiancharoen, Pawinee Doung-ngern, Somkid Kripattanapong, Soawapak Hinjoy, Thitipong Yingyong, Archawin Rojanawiwat, Aekkawat Unahalekhaka, Watcharaporn Kamjumphol, Kulsumpun Khobanan, Phimrata Leethongdee, Narisorn Lorchirachoonkul, Suwimon Khusuwan, Suwatthiya Siriboon, Parinya Chamnan, Amornrat Vijitleela, Traithep Fongthong, Krittiya Noiprapai, Phairam Boonyarit, Voranadda Srisuphan, Benn Sartorius, John Stelling, Paul Turner, Nicholas PJ Day, Direk Limmathurotsakul

**Affiliations:** Mahidol-Oxford Tropical Medicine Research Unit, Faculty of Tropical Medicine, Mahidol University, Bangkok, Thailand; University of Oxford, Oxford, United Kingdom; Ministry of Public Health, Nonthaburi, Thailand; Ratchaburi Hospital, Ratchaburi, Thailand; Chiangrai Prachanukroh Hospital, Chiang Rai, Thailand; Sunpasitthiprasong Hospital, Ubon Ratchathani, Thailand; Brigham and Women’s Hospital and Harvard Medical School, Boston, MA, United States; Cambodia-Oxford Medical Research Unit, Angkor Hospital for Children, Siem Reap, Cambodia

## Abstract

There are few studies comparing proportion, frequency, mortality and mortality rate of antimicrobial-resistant (AMR) bacterial infections between tertiary-care hospitals (TCHs) and secondary-care hospitals (SCHs) in low and middle-income countries (LMICs) to inform infection control strategies. We evaluated bloodstream infections (BSIs) from 2012 to 2015 in 15 TCHs and 34 SCHs in Thailand. There were differences in the proportions (%) of BSI caused by AMR strains for some pathogens between TCHs and SCHs. Of 19,665 patients with AMR BSI, 6,746 (34.3%) died. Among patients with AMR BSI, there were no or minimal differences in mortality proportion for all AMR pathogens between TCHs and SCHs. However, the frequency and mortality rates of AMR BSI were considerably higher in TCHs for most pathogens. For example, the mortality rate of hospital-origin carbapenem-resistant *Acinetobacter baumannii* BSI in TCHs was nearly three times higher than that in SCHs (10.2 vs. 3.6 per 100,000 patient-days at risk, mortality rate ratio 2.77; 95% confidence interval 1.71 to 4.48, p<0.0001). Targets of and resources for antimicrobial stewardship and infection control programs in LMICs may need to be tailored based on hospital type and size, as burden of AMR infections could differ by hospital setting.

## Introduction

Understanding and monitoring the burden of antimicrobial resistant (AMR) bacterial infection is important to design strategies for infection prevention and control.^1^ A recent modelling study estimated that there are 1.27 million deaths attributable to AMR infections comparing to non-AMR infections in 2019 globally.^2^ The study also highlighted the limited availability of data in LMICs,^2^ where most of the currently available data were from university hospitals and tertiary-care hospitals (TCH).^3,4^

Multiple parameters are required for monitoring and evaluating the AMR burden in hospital settings. The proportions (%) of patients with growth of AMR strains of bacterial species (over total number of patients with growth of bacterial species) are commonly used to represent AMR burden.^5,6^ However, proportions of patients with resistant infection alone cannot reflect the burden of AMR in absolute terms. For example, 10 over 20 and 50 over 100 are both 50%, but ten patients with AMR infections is a much lower burden than 50 patients with AMR infections. The frequencies of patients with AMR infections within a population during a reporting period (i.e. AMR frequencies^7^) are other important parameters. The AMR frequencies are also commonly used to monitor, evaluate and compare the AMR burden between hospitals or sentinel sites.^8-10^

AMR proportions and AMR frequencies are reported to be different by type and size of hospitals in some settings in high-income countries (HICs). In Spain, Oteo *et al*. reported that the proportion of methicillin-resistant *Staphylococcus aureus* (MRSA) is higher in hospitals with >500 beds than in those with <500 beds.^11^ In Germany, Said *et al*. reported that the proportion of carbapenem-resistant *Acinetobacter baumannii* (CRAB) is higher in TCH and secondary-care hospitals (SCH) compared to outpatient clinics.^12^ In the U.S., Gandra *et al*. reported that the proportion of AMR infections is not different between TCH and small community hospitals.^13^ The point prevalence survey of healthcare-associated infections (HAI) in European acute care hospitals shows that HAI prevalence is highest in hospitals with ≥650 beds and lowest in those with <200 beds,^14^ suggesting that frequency of HAI (per patients who were admitted to the hospital) is associated with hospital size.

We recently developed the AutoMated tool for Antimicrobial resistance Surveillance System (AMASS), an offline application to generate standardized AMR surveillance reports from routinely available microbiology and hospital data files, and independently tested the application in seven hospitals in seven countries.^15^ The automatically generated reports stratify infections into community-origin and hospital-origin based on the recommendations of World Health Organization Global Antimicrobial Resistance and Use Surveillance System (WHO GLASS), and provide additional metrics on mortality involving AMR and non-AMR bloodstream infections (BSI).^15^ Collaborating with Ministry of Public Health (MoPH) Thailand, we previously obtained and analysed microbiology and hospital admission data files of 60 hospitals from 2012 to 2015 in Thailand, and reported the burden of melioidosis, an infectious disease caused by *Burkholderia pseudomallei*, in Thailand.^16^

The aim of this study was to examine the burden of AMR BSI in TCHs and compare that with SCHs using AMASS^15^ on the microbiology and hospital admission data from 2012 and 2015 in Thailand.

## METHODS

### Study setting

In 2012, Thailand had a population of 64.4 million, consisted of 77 provinces, and covered 513,120 km^2^. The country is divided into six primary administrative geographical regions (provinces), comprising Northeast, North, East, West, South, and Central. In each province, there is at least one SCH or TCH,^17^ equipped with a microbiology laboratory capable of performing bacterial culture using standard methodologies for bacterial identification and susceptibility testing provided by the Bureau of Laboratory Quality and Standards, MoPH, Thailand.^18^

### Study design

We conducted a retrospective, multicentre surveillance study of all SCH and TCH hospitals in Thailand. From the hospitals that agreed to participate, data were collected from microbiology and hospital admission data files between January 2012 and December 2015 as previously described.^16^ Each data set was analysed using the AMASS v2.0,^19^ and patient hospital number (HN) was used as a record linkage between the two data files of each hospital. The AMASS analysed the data and generated reports based on the recommendation of WHO GLASS.^20^ A deduplication process was automatically conducted in which only the first isolate of a species per patient per specimen type per survey period was included in the report.^15^ The AMASS v2.0 included an additional report on notifiable bacterial diseases (Annex A) and blood culture contamination rate (Annex B) (Supplementary file 1). The statistics in the AMR surveillance reports (in PDF and CSV format) were then extracted for analysis.

For AMR infections, we analysed the following organisms: CRAB and carbapenem-resistant *Pseudomonas aeruginosa* (CRPA), third-generation cephalosporin-resistant *Escherichia coli* (3GCREC) and *Klebsiella pneumoniae* (3GCRKP), carbapenem-resistant *E. coli* (CREC), carbapenem-resistant *K. pneumoniae* (CRKP) and MRSA which are in the WHO GLASS list of priority AMR bacteria^21^ and are of locally importance. Only blood culture isolates were included in the analysis.

### Definitions

AMR BSI is defined as a case of infection in patients with blood culture positive for CRAB, CRPA, 3GCREC, 3GCRKP, CREC, CRKP or MRSA. Non-AMR BSI is defined as cases of infection in patients who had blood culture positive for carbapenem-susceptible *A. baumannii* (CSACI), carbapenem-susceptible *P. aeruginosa* (CSPA), third-generation cephalosporin-susceptible *E. coli* (3GCSEC) or *K. pneumoniae* (3GCSKP), or methicillin-susceptible *S. aureus* (MSSA).

Community-origin BSI was defined for patients with first positive blood specimens in the hospital taken within the first two calendar days of admission with calendar day one equal to the day of admission.^20^ Patients with first positive blood specimens taken after the first two calendar days were categorized as cases of hospital-origin BSI.

The proportion of AMR (%) was calculated as the percentage of patients with new AMR BSI over all patients with new BSIs for each pathogen of interest during the reporting period.^19^ The frequency of AMR BSI for each pathogen of interest was calculated as the total number of new patients with AMR BSI during the reporting period per 100,000 admissions (for community-origin BSI), per 100,000 patient-days at risk (for hospital-origin BSI), and per 100,000 tested population (for community-origin and hospital-origin BSI). In-hospital mortality (%) of AMR BSI for each pathogen of interest was calculated as the percentage of patients with new AMR BSI who died in the hospitals. Mortality rates for AMR BSI for each pathogen of interest were calculated as the total number of patients with new AMR BSI who died in the hospitals during the admission with AMR BSI per 100,000 admissions (for community-origin BSI) and per 100,000 patient-days at risk (for hospital-origin BSI).

In the AMASS 2.0, blood culture contamination is defined as isolation of one or more common commensal organisms; including *Arcanobacterium* spp., *Arthrobacter* spp., *Bacillus* spp. (except *B. anthracis*), *Brevibacillus* spp., *Brevibacterium* spp., *Cellulomonas* spp., *Cellulosimicrobium* spp., *Corynebacterium* spp. (except *C. diphtheriae, C. jeikeium, C. pseudotuberculosis, C. striatum, C. ulcerans*, and *C. urealyticum*), *Cutibacterium* spp., *Dermabacter* spp., *Dermacoccus* spp., *Diphtheroids* spp., *Exiguobacterium* spp., *Geobacillus* spp., *Helcobacillus* spp., *Janibacter* spp., *Knoellia* spp., *Kocuria* spp., *Kytococcus* spp., *Leifsonia* spp., *Microbacterium* spp., *Micrococcus* spp., *Nesterenkonia* spp., *Paenibacillus* spp., *Propionibacterium* spp., *Pseudoclavibacter* spp., *Staphylococcus* spp. (except *S. aureus* and *S lugdunensis*), *Trueperella* spp., *Virgibacillus* spp., and Viridans group *Streptococci*.^18^ The blood culture contamination rate is defined as the ratio of the number of blood cultures with common commensal organisms over the total number of blood cultures.

### Statistical analysis

We compared AMR proportions using Chi-square or Fisher’s exact test when small samples (<5 observations in any one strata) and measurements from continuous variables using the Krustal-Wallis test. We also estimated the magnitude of differences in proportions and frequency of AMR BSI between SCHs and TCHs using mixed-effect logistic and Poisson regression models for patients nested within hospitals, respectively. We used STATA (version 17.0; College Station, Texas) for the final statistical analyses and R version 4.0.5 for figures.

### Data availability

The anonymous AMR surveillance reports generated from each hospital are open-access and available at https://figshare.com/s/c028f157c18a3cc06a82.

### Ethical Considerations

Ethical permission for this study was obtained from the Institute for the Development of Human Research Protection, Ministry of Public Health (IHRP 2334/2556), the Ethics Committee of the Faculty of Tropical Medicine, Mahidol University (MUTM 2014-017-01), and the Oxford Tropical Research Ethics Committee, University of Oxford (OXTREC 521-13). Written approval was given by the directors of the hospitals to use their routine hospital admission database for research. Individual consent was not sought from the patients as this was a retrospective study, and the Ethical and Scientific Review Committees approved the process.

## RESULTS

### Baseline demographics

Of the 96 Thai hospitals approached (28 TCHs and 68 SCHs in 2012, 95 (99%) agreed to participate in the study. Twenty-five hospitals (26%) were excluded because either the microbiology or hospital admission database was not available. Next, twenty-one hospitals were excluded because the antimicrobial susceptibility tests results were not available. Forty-nine hospitals were included in the analysis. A total of 35 hospitals (71%) had four years (from 2012 to 2015) of data available for analysis, four hospitals (8%) had three years, four hospitals (8%) had two years, and six hospitals (12%) had one year of data for analysis (Supplementary file 2).

Of 49 hospitals included in this study, 15 (31%) and 34 (69%) were TCHs and SCHs respectively (Figure 1). The median bed number was 672 (range 522-1,000) in TCHs and 335 (range 150-549) in SCHs (p=0.0001). The median number of hospital admissions per year was 48,836 (range 30,409-98,428) in TCHs and 25,827 (range 7,221-62201) in SCHs (p=0.0001). The total number of admissions was 3,134,815 in TCHs and 2,867,762 in SCHs. The blood culture utilization rate was slightly higher in TCHs compared to SCHs (median blood culture utilization rate 69 vs. 60 per 1,000 patient-days, p=0.12). The blood culture contamination rate was not different between TCHs and SCHs (median blood culture contamination rates were 4.1% and 3.6%, respectively, p=0.94). The in-hospital mortality among all patients admitted to TCHs was higher than those to SCHs (median in-hospital mortality 3.7% vs. 2.9%, respectively, p=0.05).

**Figure 1.**
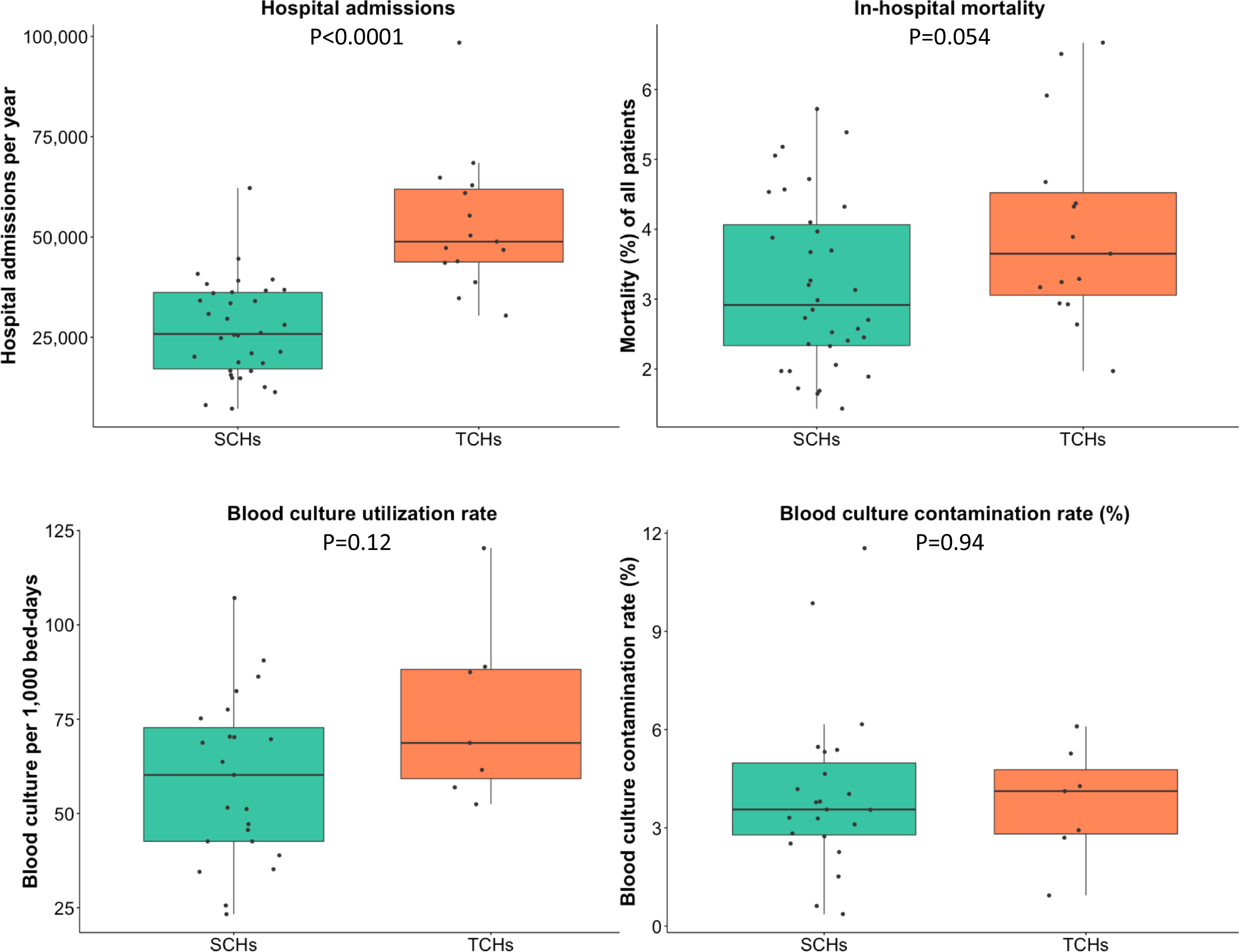
Baseline demographics of 15 tertiary-care hospitals and 34 secondary-care hospitals in Thailand. Each black dot represents the value reported by each hospital. Comparison was made using the Kruskal-Wallis test. The unit of analyses was a hospital. Blood culture (BC) utilization rate (per 1,000 bed-days) and BC contamination rate (%) were estimated from 7 TCHs and 23 SCHs of which the microbiology data obtained included the culture results of “no growth”.

### Proportion of AMR BSI

For community-origin BSI, there were differences in the proportions (%) of BSI being caused by AMR strains for some pathogens between TCHs and SCHs. The proportions of CRAB (36.9% vs. 25.0%, p=0.041), 3GCREC (37.7% vs. 31.2%, p=0.020) and 3GCRKP (24.4% vs. 19.2%, p=0.053) were higher in TCHs than those in SCHs (Figure 2A and Supplementary File 3). The proportions of CRPA (18.4% vs. 17.1%), CREC (0.9% vs. 1.6%) and CRKP (2.3% vs. 2.7%) and MRSA (11.7% vs. 11.2%) were not different between TCHs and SCHs (all p>0.20).

**Figure 2.**
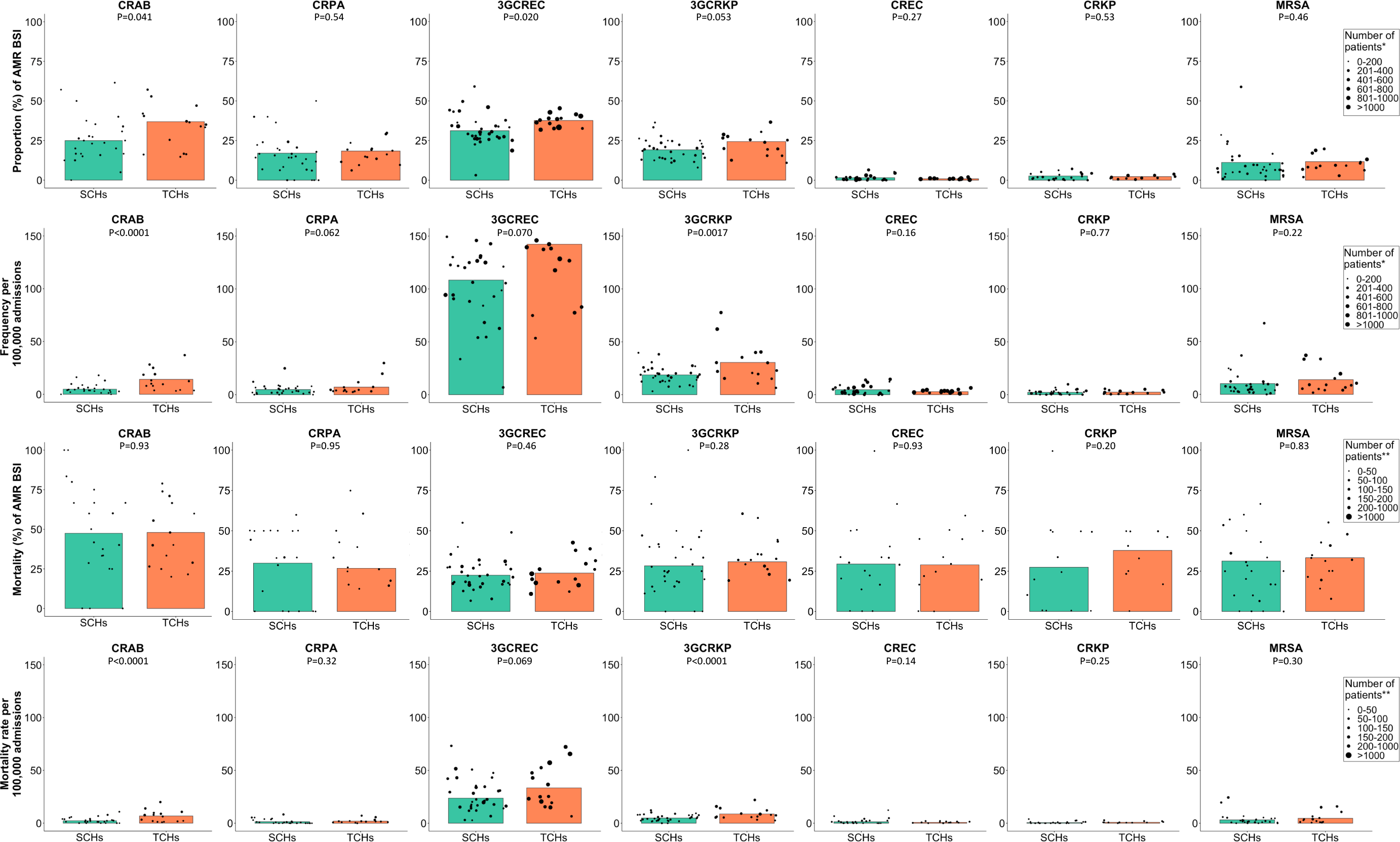
Proportion (%), frequency (number of patients per 100,000 admissions), mortality (%) and mortality rate (number of deaths per 100,000 admissions) of community-origin AMR BSI in 15 tertiary-care hospitals and 34 secondary-care hospitals in Thailand. Each black dot represents the value reported by each hospital. The sizes of the black dots are based on the total number of patients with blood culture positive for the bacterial species (row 1) and the total number of patients with blood culture positive for the AMR pathogens (row 2-4).

For hospital-origin BSI, the proportions of AMR BSI for all pathogens, including CRAB (75.5% vs. 68.3%), CRPA (36.7% vs. 36.7%), 3GCREC (53.9% vs. 53.5%), 3GCRKP (63.7% vs. 57.3%), CREC (3.0% vs. 3.2%), CRKP (9.2% vs. 8.6%) and MRSA (37.4% vs. 28.0%), were not different between TCHs and SCHs (all p>0.10; Figure 3A and Supplementary File 3).

**Figure 3.**
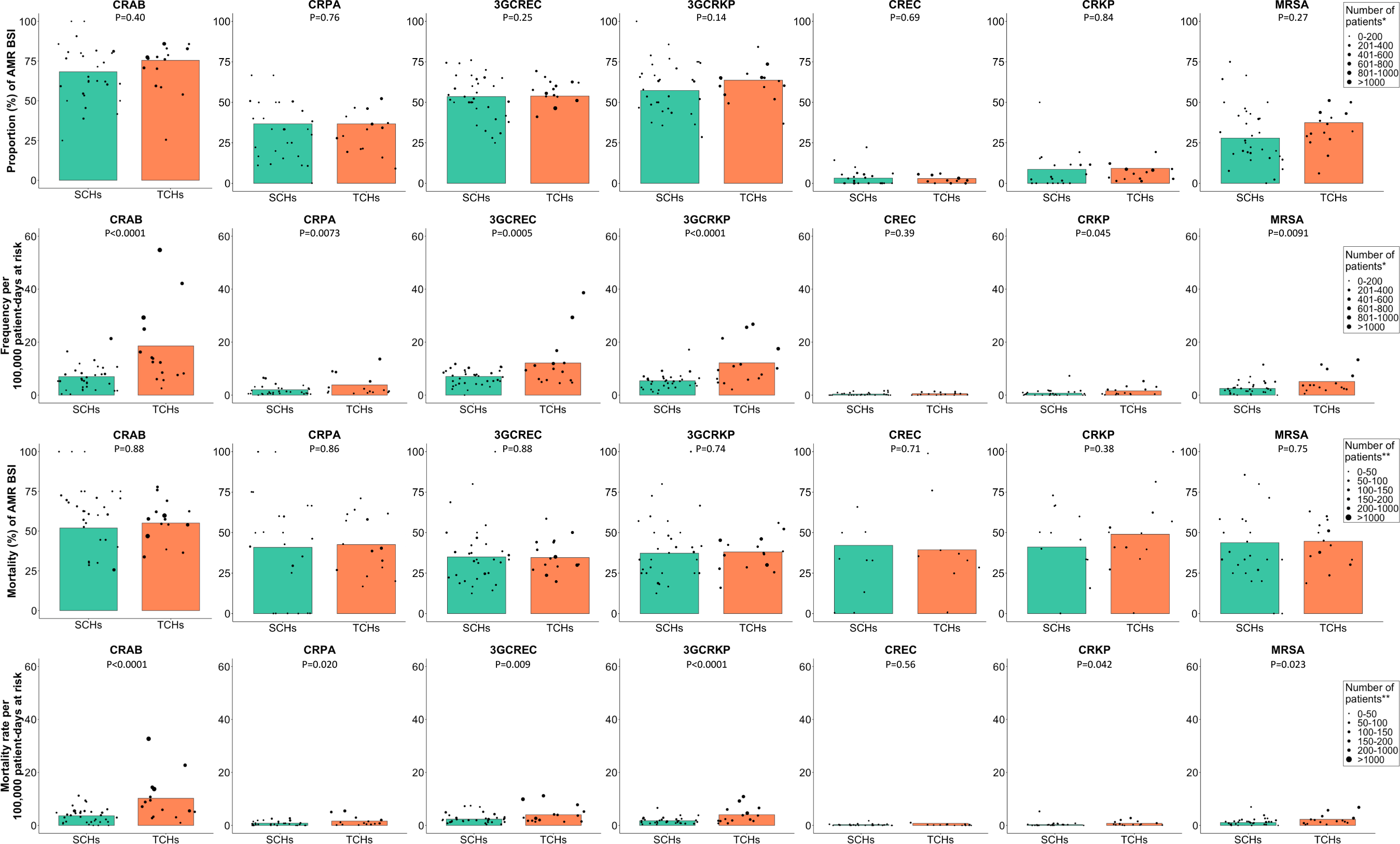
Proportion (%), frequency (number of patients per 100,000 patient-days at risk), mortality (%) and mortality rate (number of deaths per 100,000 patient-days at risk) of hospital-origin AMR BSI in 15 tertiary-care hospitals and 34 secondary-care hospitals and in Thailand. Each black dot represents the value reported by each hospital. The sizes of the black dots are based on the total number of patients with blood culture positive for the bacterial species (row 1) and the total number of patients with blood culture positive for the AMR pathogens (row 2-4).

### Frequency of AMR BSI

We next calculated the frequency of AMR BSI in SCHs and TCHs. For community-origin BSI, of all pathogens under evaluation, 3GCREC had the single highest frequency of AMR BSI per 100,000 admissions in both TCHs and SCHs (Figure 2B and Supplementary file 4). The frequencies per 100,000 admissions of community-origin BSI caused by CRAB (14.2 vs. 4.8, p<0.0001), CRPA (7.1 vs. 4.9, p=0.062), 3GCREC (142.3 vs. 108.4, p=0.070) and 3GCRKP (30.5 vs. 18.8, p=0.0017) in TCHs were relatively higher than those in SCHs. The frequencies of CREC (2.9 vs. 4.7), CRKP (2.4 vs 2.2) and MRSA (14.0 vs. 10.1) were not different between TCHs and SCHs (all p>0.10).

Overall, the number of patient-days at risk of hospital-origin BSI included in the analysis was 12,341,585 in TCHs and 9,988,198 in SCHs. For hospital-origin BSIs, of all pathogens under evaluation, CRAB, 3GCRE and 3GCRKP were the top three organisms with the highest frequency in both TCHs and SCHs (figure 3B and Supplementary file 5). Strikingly, the frequency of hospital-origin BSI per 100,000 patient-days at risk in TCHs was about twice that in SCHs for most of the pathogens under evaluation, including CRAB (18.6 vs. 7.0, incidence rate ratio [IRR] 2.77; 95%CI 1.72-4.43, p<0.0001), CRPA (3.8 vs. 2.0, IRR 2.14; 95%CI 1.23-3.74, p=0.0073), 3GCREC (12.1 vs. 7.0, IRR 1.80; 95%CI 1.29-2.50, p=0.0005), 3GCRKP (12.2 vs. 5.4, IRR 2.23; 95%CI 1.57-3.17, p<0.0001), CRKP (1.6 vs. 0.7, IRR 2.10; 95%CI 1.02-4.35, p=0.045) and MRSA (5.1 vs. 2.5, IRR 1.88; 95%CI 1.17-3.01, p=0.0091), except CREC (0.5 vs. 0.4, p=0.39).

Just under half of the hospitals included in this study could only provide data on blood cultures positive for. The frequencies of AMR BSI per 100,000 tested patients were calculated for 7 (47%) TCHs and 23 (68%) SCHs, which could provide results for blood cultures without growth (Supplementary File 6). Similar findings of difference between TCHs and SCHs were also observed, but wider 95%CI and larger p values were observed.

### Mortality (%) of patients with AMR BSI

Of 19,665 patients with AMR BSI caused by pathogens under evaluation, 6,746 (34.3%) died. For both community-origin and hospital-origin BSIs, of all pathogens under evaluation, mortality of patients with AMR BSI caused by CRAB was higher than AMR BSI caused by other pathogens (Figure 2C and 3C, and Supplementary File 7). For both community-origin BSI and hospital-origin BSI, there were no or minimal differences in the mortality (%) of patients with AMR BSI caused by all pathogens between TCHs and SCHs (all p>0.20).

### Mortality rate for AMR BSI

We next calculated mortality rate for AMR BSI in SCHs and TCHs. For community-origin BSI, of all pathogens under evaluation, 3GCREC had the single highest mortality rate of AMR BSI per 100,000 admissions in both TCHs and SCHs (Figure 2D and Supplementary file 8). The mortality rate per 100,000 admissions of community-origin BSI caused by CRAB, 3GCREC and 3GCRKP in TCHs was relatively higher than in SCHs (6.8 vs. 2.3; mortality rate ratio [MRR] 2.92, 95%CI: 1.65-5.19, p<0.0001, 33.4 vs. 23.7; MRR 1.40, 95%CI 0.97-2.01, p=0.069 and 8.8 vs. 4.9; MRR 1.79, 95%CI 1.28-2.52, p<0.0001, respectively). There were only minor differences in the mortality rate per 100,000 admissions of community-origin BSI being caused by CRPA, CREC, CRKP and MRSA between TCHs and SCHs (all p>0.10).

For hospital-origin BSI, of all pathogens under evaluation, CRAB had the single highest mortality rate per 100,000 patient-days at risk in both TCHs and SCHs (Figure 3D and Supplementary file 9). Strikingly, the mortality rate per 100,000 patient-days at risk of hospital-origin BSI caused by most of pathogens under evaluation was also about two to nearly three times those in SCHs; including CRAB (10.2 vs. 3.6; MRR 2.77, 95%CI 1.71-4.48, p<0.0001), CRPA (1.6 vs. 0.8, p=0.020), 3GCREC (4.0 vs. 2.4, p=0.009), 3GCRKP (4.0 vs. 1.8, p<0.0001), CRKP (0.8 vs. 0.3, p=0.042) and MRSA (2.3 vs. 1.1, p=0.023), though not for CREC (0.2 vs. 0.2, p=0.56).

### Notifiable bacterial diseases

Utilizing microbiology data and hospital admission data, we also calculated total number and in-hospital mortality of patients with culture-confirmed notifiable bacterial diseases indicated in the National Notifiable Disease Surveillance system (Report 506) of Thailand,^22^ in the 49 hospitals from 2012 to 2015 (Supplementary File 10). The disease with the highest total number of cases was non-typhoidal *Salmonella* (NTS) infection (n=11,264 patients), followed by melioidosis (an infection caused by *Burkholderia pseudomallei*; n=6,164 patients) and *Vibrio* spp. infections (n=2,143 patients). The disease with the highest total number of in-hospital deaths was melioidosis (n=1,524 patients), followed by NTS infection (n=1,005 deaths), *Vibrio* spp. infection (n=172 deaths), *Streptococcus suis* infection (n=85 deaths), *Corynebacterium diphtheriae* infection (n=9 deaths), *Shigella* spp. infection (n=7 deaths), *Salmonella enterica* serovar Paratyphi infection (n=4 deaths), *S. enterica* serovar Typhi infection (n=3 deaths), and *Neisseria meningitidis* infection (n=3 deaths). None of 60 and 4 patients with culture-confirmed *N. gonorrhoeae* infection and *Brucella* spp. infection died in the hospital.

## DISCUSSION

This study compared proportions, frequencies, mortality and mortality rate of AMR BSI between TCHs and SCHs in a LMIC. We show that the frequency and mortality rates of AMR BSI were considerably higher in TCHs than those in SCHs. The results support the needs to design antimicrobial stewardship (AMS) and infection prevention and control (IPC) in LMICs base on hospital size and type.

Our study highlights the capability of hospitals and national authorities in LMICs to estimate frequencies as a crucial parameter to monitor the effectiveness of AMR interventions. For example, effectiveness of a nationwide intervention in Israel is shown by the reduction in frequency of nosocomial carbapenem-resistant *Enterobacteriaceae* infections from 55.5 to 11.7 cases per 100,000 patient-days.^23^ An 80% reduction in MRSA BSI in England (defined as reported cases per 100,000 population per year and as reported cases per 100,000 bed-days) after a major public health infection prevention campaign also demonstrates the potential impact of coordinated interventions.^24,25^

The higher proportion (%) and frequencies of community-origin AMR BSI for CRAB, 3GCREC and 3GCRKP in TCHs compared to those in SCHs could be due to higher proportion of healthcare-associated infections in TCHs. It is likely that TCHs have a higher proportion of patients who are transferred from other hospitals (including from SCHs), who are receiving health care at end-stage renal facilities or long-term care facilities, and who are recently discharged from the hospitals. Routine hospital admission data used in our study could not identify those conditions; therefore, those patients with BSI were categorized as community-origin BSIs in our reports. These findings suggest that TCHs may need to strengthen AMS and IPC, particularly on new inpatients who are at high risk of healthcare-associated infections.

No or minimal difference in proportion (%) of hospital-origin AMR BSI between TCHs and SCHs could be because the AMR prevalence in a country is likely driven by the contagion of AMR organisms within and between hospitals.^26^ Although it is possible that higher use of antibiotics in TCHs may drive the emergence of AMR organisms (e.g. emergence of CREC and CRKP) and lead to the higher proportion of AMR BSI compared to those in SCHs,^27^ that was not observed in our setting during the study period.

The strikingly higher frequencies of hospital-origin AMR BSI in TCHs than those in SCHs are likely caused by higher proportion of patients who had severe conditions or compromised immune systems, or required complex surgery, prolonged intubation or urinary catheters.^11,14^ The proportion of ICU beds in TCHs is also higher. Those conditions are likely driving both AMR and non-AMR hospital-acquired infections in our setting as shown by no or minimal difference in proportion of hospital-origin AMR BSI between TCHs and SCHs.

No or minimal difference in mortality (%) of AMR BSI between TCHs and SCHs means that once a patient has AMR BSI, the patient has a comparable chance of death. This could be because care and antibiotics to be used against AMR BSI are not different between TCHs and SCHs in Thailand.

The higher mortality rate of hospital-origin AMR BSI in TCHs than those in SCHs is, therefore, caused by higher frequencies of AMR BSI. These findings suggest that healthcare workers in TCHs will need to strengthen AMS and IPC, particularly among those are at high risk of hospital-acquired infections (HAI), more than those in SCHs. This is also because patients in TCHs are likely to be more complex than those in SCHs, and, as such, are at higher risk of HAI than those in SCHs.

For hospital-origin AMR BSI in SCHs, although CRAB, 3GCREC and 3GCRKP are the top three organisms with the highest frequencies, CRAB has the highest mortality rate due to the higher mortality (%) of patients with CRAB BSI compared to those with 3GCREC BSI and 3GCRKP BSI.

Our findings of high frequencies and deaths of nontyphoidal *Salmonella* disease,^28,29^ melioidosis,^16,30^ *S. suis* infection,^31,32^ and *Vibrio* spp. infection^33,34^ are consistent with previous findings. Our study demonstrates that national statistics on multiple national notifiable bacterial diseases in LMICs could be improved by integrating information from readily available databases. The strength of this study is that routine data used to compare AMR burdens between different hospital settings are from multiple sites in Thailand. Moreover, we have shown that the use of an automated surveillance system can readily generate useful statistics to understand AMR within a hospital and between hospitals in a country, and this empowers collaborative work and analyses across different settings both nationally and globally. The collaborative effort is essential to inform global burdens of AMR, which in turn are important statistics to support public health strategies to control spread of AMR and set priorities in resource allocation locally.

Our study has some limitations. Firstly, the findings from the study may not be generalisable to all LMICs. The differences in proportions, frequencies, mortality and mortality rates of AMR BSI in SCHs and TCHs are confounded by many factors such as difference in true susceptibility profiles of pathogenic organisms in the settings, patient characteristics, diagnostic stewardship (particularly blood culture utilization^35,36^) and patient management (including AMS and IPC). Secondly, the sample size of SCHs and TCHs included in this study is small, and this may have limited the power to detect differences for organisms that are less predominate such as CRKP in the study setting. Thirdly, in this study we only included patients who were hospitalised and in-hospital mortality. Patients who had blood cultures taken either at community hospitals or the study hospitals but not hospitalised at the study hospitals were not included in the analysis. Fourthly, the mortality and mortality rate associated with AMR BSI reported could be underestimated because some people, in the study area, preferred to die at home and were discharged against advice.

In conclusion, the burden of AMR infections in TCHs is higher than that in SCHs in Thailand. This is likely occurring in other LMICs. Our results support the importance of tailoring infection control strategies based on hospital size and type.

## Data Availability

All data produced in the present work are contained in the manuscript

https://figshare.com/s/c028f157c18a3cc06a82

## Acknowledgement

We gratefully acknowledge the directors, epidemiological and laboratory team of general and regional hospitals for providing microbiological and hospital admission data and their administrative supports. The general and regional hospital network are comprised of Chorkaew Yangyuen (Samutprakarn Hospital), Aphinya Singkhongsin, Chanchira Chaichaem (Pranangklao Hospital), Phkaiwan Kropsanit (Pathumthani Hospital), Winai Suphapphot, Chaiwat Khaokaeo (Sena Hospital), Sasi Sichot (Phra Na Khon Sri Ayutthaya Hospital), Ratri Chalaemphak, Benchawan Khaisongkhram (Angthong Hospital), Praphon Chinthanu, Phutthakhun Wongsuwan (Banmi Hospital), Pritsana wongnoi (Pranarai Maharaj Hospital), Witthaya Yotngoen (Singburi Hospital), Khongsak Sueachoi (Inburi Hospital), Pranom Chantharat (Jainad Narendra Hospital), Sangsan Sinbamrung, Duangkamon Chiratrachu (Phra Phutthabat Hospital), Waranya Sichanta (Saraburi Hospital), Panatda Thipruecha (Chonburi Hospital), Piyaphatcha Phongprasoet, Panatda Inphrom (Rayong Hospital), Pakkawi Siphueak, Ratchani Thamchamrat (Prapokklao Hospital), Phuangphikun PhonPrasit (Trat Hospital), Bunga Chanlee (BuddhaSothorn Hospital), Wiphawadi Dongchan (Chao Phya Abhaibhubejhr Hospital), Haruethai Khunothai (Nakhonnayok Hospital), Atchara Ampere (Somdet Phrayupharacha SaKaeo), Saifon Sutchai, Prayut Kaeomalang, Nonglak Prayunsoet (Maharat Nakhon Ratchasima Hospital), Ratana Chiracharuporn (Buriram Hospital), Suriya Senthong (Surin Hospital), Sunthon Romniyaphet (Sisaket Hospital), Jintana Kanchanabat, Praweennuch Watanachaiprasert, Thanasith Sananmuang (Sunpasitthiprasong Hospital), Somphon Chankaeo (Yasothon Hospital), Wiraphon Khwamman (Chaiyaphum Hospital), Kraison Bunsam, Phonnatcha Katiwong (Amnat Charoen Hospital), Kanyaphak Phanchampa (Buengkan Hospital), Sutthiphong Phonbun (Nongbualamphu Hospital),Marisa Uton, Thitiphan Khunphu (Sirindhorn Hospital of Khon Kaen Province), Kriangkri Kongsuk, Ritthikorn DongLuang (Khon Kaen Hospital), Suthep Thipsawang, Kochnipa Kwawong, Nida Thanaphatphairot (UdonThani Hospital), Suphattra Likrachang, Kirana Phakdiburikun (Loei Hospital), Suphakon Saenthamphon (Nongkhai Hospital), Suchitra Nasingkhan, Kanchanaphon Taratai (Mahasarakham Hospital), Witthaya Ratmaet (Roiet Hospital), Natthasorn Chawaninthawisut (Kalasin Hospital), Phuwanat PhothiChai (Sakonnakhon Hospital), Phinthip Saiklang (Nakhonphanom Hospital), Yutphon Mankhong, Yothin Tairayawong (Mukdahan Hospital), Warawan Inthip (Nakornping Hospital), Dr. Thiraphong Tatiyaphonkunthiraphong (Lamphun Hospital), Thirin Ketwichit (Lampang Hospital), Yaowalak ChanDaeng (Uttaradit Hospital), Phana Thatsanawaythit, Itsareeya Boonrat (Phrae Hospital), Sopha Itsaranarongphan (Nan Hospital), Sanong Chaisue, Phanarat Phuangmali (ChiangKham Hospital), Chirawan Sithongphim (Phayao Hospital), Satorn Charatdamrongwat (Chiangrai Prachanukroh Hospital), Jintana Phothip, Duangdi Chomphu (Srisangwan Hospital), Ladda Raden (Sawanpracharak Hospital), Phitya Hema, Kanthika Ocharot, Mongkhon Uising (Uthaithani Hospital), Narong Mahayot (Kampangpetch Hospital), Onraphin Thiwai (Maesot Hospital), Preeyada Triprawat (Somdejprajaotaksin Maharaj Hospital), Kreangkrai Chatsut, Yuppharet Kaewprasern (Srisangworn Sukhothai Hospital), Meena Nakhon (Sukhothai Hospital), Aphinya Innoi, Thoranin Rakthanabodee (Buddhachinaraj hospital), Siwaphorn Phongchin (Phichit Hospital), Jintana Phonphraram (Phetchabun Hospital), Somphon Niamlang (Damnoensaduak Hospital), Thanya Surakhamsang (Banpong Hospital), Thanyalak Borirak (Photharam Hospital), Nopphon Siangchin (Ratchaburi Hospital), Ratchani Watthanayaem, Suwan Manutchan (Pahonpol Payuha Sena Hospital), Ekachai Photnanthawong (Makarak Hospital), Dr. Pornsak Thirathonbun (SomdejPrasangkharach17 Hospital), Narong Wongkanha (Chaophraya Yommarat Hospital), Pitchayakhanid Yaemsoun, Pongphon Roeknaowarat (Nakhonpathom Hospital), Witthaya Sithong (Samutsakorn Hospital), Nonsi Sonthiyat (Kratumban Hospital), Narongchai SiamPhairi (Somdej Prabuddha Lertla Hospital), Chanthana Kalanuwat (Prajomklao Hospital), Saran Songsaeng (HuaHin Hospital), Chirawan Bunchusi (Maharaj Nakhonsithammarat Hospital), Chitchanin Niyomthai, Chutima Phayayam (Krabi Hospital), Boribun Chensamut (Takuapa Hospital), Aphisit Suwannarat, Suwanni Khwankhao, Phityaporn Chunchu (Phang Nga Hospital), Phatcharin Yatraksa (Vachira Phuket Hospital), Jittima Thongnak (Koh Samui Hospital), Ratchanok Withunphan (Suratthani Hospital), Chuenkhwan Kaeowichit, Phimnisa Phet (Ranong Hospital), Kritsanee Wichitakun (Chumporn Ketudomsak Hospital), Sakda Khaophong (Songkla Hospital), Wichian Patangkaro, Nattawan Chanmueang (Hat Yai Hospital), Nittaya Sakunsanti (Satun Hospital), Suriwan Phaksuphara, Umaporn Sina (Trang Hospital), Witthaya Wunchum (Phatthalung Hospital), Sirinthon Wongyoksuriya (Pattani Hospital), Mawin Deae, Ananni Sama (Betong Hospital), Wichai Wanmueang, Suphattra Mahachot (Yala Hospital), Arun Phutkaeo, Yukalipli Kaseng, Khodiyo Yamai (Narathiwat Ratchanakarin Hospital), Praphai Krirat, Naruemon Bunsiri (Sungai Kolok Hospital). We thank Saman Sayumphuruchinan (ED, MoPH), Wanwisa Khammak (Strategy and Planning Division, MoPH), Sittikorn Rongsumlee (MORU) and Prapass Wanapinij (MORU) for administrative and data management supports.

The study was supported by the DDC, MoPH, Thailand, and Defense Threat Reduction Agency (DTRA), U.S.. This research was funded in part by the Wellcome Trust [224681/Z/21/Z and Wellcome Trust Institutional Translational Partnership Award-MORU]. BS is supported by a grant from the UK Department of Health and Social Care using UK aid funding managed by the Fleming Fund (R52354 CN001). For the purpose of Open Access, the author has applied a CC BY public copyright licence to any Author Accepted Manuscript version arising from this submission

## Notes

### Competing Interest Statement

The authors have declared no competing interest.

### Author Declarations

The study used ONLY openly available human data that were originally located at: https://figshare.com/s/c028f157c18a3cc06a82. Ethical permission for this study was obtained from the Institute for the Development of Human Research Protection, Ministry of Public Health (IHRP 2334/2556), the Ethics Committee of the Faculty of Tropical Medicine, Mahidol University (MUTM 2014-017-01), and the Oxford Tropical Research Ethics Committee, University of Oxford (OXTREC 521-13). Written approval was given by the directors of the hospitals to use their routine hospital admission database for research.

### Summary of Updates

This version of the manuscript has been revised to updated data from anonymous hospital no. 17. The microbiology data files of all participating hospitals reported species of Acinetobacter in details (e.g. A. baumannii, Acinetobacter baumannii-calcoaceticus complex, A. nosocomialis, etc), except the microbiology data file of anonymous hospital no. 17, which reported all Acinetobacter as Acinetobacter spp.. In the current version of the manuscript, we assumed that all isolates reported as Acinetobacter baumannii-calcoaceticus complex reported in all participating hospitals and all isolates reported as Acinetobacter spp. in the anonymous hospital no. 17 were A. baumanii. The revision has not changed the conclusion. The data of all other pathogens are the same as that in the previous version.

