## Supplementary materials for "Higher Frequency and Mortality Rate of Antimicrobial-resistant Bloodstream Infections in Tertiary-care Hospitals Compared with Secondary-care Hospitals in Thailand"

**Supplementary File 1. Supplementary text**

### METHODS

#### AMASS 2.0

AMASS v2.0 was modified from AMASS v1.1.[1] The AMASS v2.0 included an additional report on notifiable bacterial diseases (Annex A) and data indicators (Annex B). Data indicators reported included blood culture contamination rate and proportion of isolates with infrequent phenotypes or potential errors in AST results.

List of notifiable bacterial diseases was derived from bacterial diseases in the National Notifiable Disease Surveillance system (Report 506) of Thailand.[2] The bacterial diseases included were *Burkholderia pseudomallei* infection (i.e. melioidosis), *Brucella* spp. infection, *Corynebacterium diphtheriae* infection, *Neisseria gonorrhoeae* infection, *Neisseria meningitidis* infection, *Non-typhoidal Salmonella* spp. infection, *Salmonella enterica* serovar Paratyphi infection, *Salmonella enterica* serovar Typhi infection, *Shigella* spp. infection, *Streptococcus suis* and *Vibrio* spp. infection. Each disease was defined based on culture positivity for the organism in any clinical specimen.

Blood culture contamination is defined as isolation of one or more common commensal organisms; including *Arcanobacterium* spp., *Arthrobacter* spp., *Bacillus* spp. (except *B. anthracis*), *Brevibacillus* spp., *Brevibacterium* spp., *Cellulomonas* spp., *Cellulosimicrobium* spp., *Corynebacterium* spp. (except *C. diphtheriae*, *C. jeikeium*, *C. pseudotuberculosis*, *C. striatum*, *C. ulcerans*, and *C. urealyticum*), *Cutibacterium* spp., *Dermabacter* spp., *Dermacoccus* spp., *Diphtheroids* spp., *Exiguobacterium* spp., *Geobacillus* spp., *Helcobacillus* spp., *Janibacter* spp., *Knoellia* spp., *Kocuria* spp., *Kytococcus* spp., *Leifsonia* spp., *Microbacterium* spp., *Micrococcus* spp., *Nesterenkonia* spp., *Paenibacillus* spp., *Propionibacterium* spp., *Pseudoclavibacter* spp., *Staphylococcus* spp. (except *S. aureus* and *S lugdunensis*), *Trueperella* spp., *Virgibacillus* spp., and Viridans group *Streptococci*.[3] The blood culture contamination rate is defined as the ratio of the number of blood culture contamination per number of total blood cultures.

Isolates with infrequent phenotypes or potential errors in AST results are defined using the isolate alert list of WHONET[4] and European Committee on Antimicrobial Susceptibility Testing (EUCAST).[5] For example, *Enterobacteriaceae* isolates with antimicrobial susceptibility test (AST) results returned as resistant to amikacin and susceptible to gentamicin would be considered as potential errors in AST results. AMASS v2.0 is open-access and available at (https://doi.org/10.6084/m9.figshare.19771186).

#### Statistical analyses

P-values were given to two significant figures, and no longer than 4 decimal places (e.g. p<0.0001). Box and dot plots were used to illustrate the distribution of hospital admissions, in-hospital mortality, blood culture utilization rate and blood culture contamination rate reported by each hospital. Bar and dot plots were used to illustrate the distribution of proportion, frequency, mortality and mortality rate of AMR BSI reported by each hospital. To represent the total burden, the bars in the figure 2 and 3 were generated by calculating the total number of numerators and denominators from all hospitals (Supplementary file 3-6).

The microbiology data files of all participating hospitals reported species of *Acinetobacter* in details (e.g. *A. baumannii*, *Acinetobacter baumannii-calcoaceticus* complex, *A. nosocomialis*, etc), except the microbiology data file of anonymous hospital no. 17, which reported all *Acinetobacter* as *Acinetobacter* spp.. In the analysis of this study, we assumed that all isolates reported as *Acinetobacter baumannii-calcoaceticus* complex reported in all participating hospitals and all isolates reported as *Acinetobacter* spp. in the anonymous hospital no. 17 were *A. baumanii.*

**Supplementary File 2. Data availability of 49 hospitals included in the analysis**

| Hospital | Data availability (year) | Hospital bed number | Hospital type* |
|---|---|---|---|
| Anonymous hospital No. 1 | 2015 | 446 | SCH |
| Anonymous hospital No. 2 | 2012-2015 | 312 | SCH |
| Anonymous hospital No. 3 | 2012-2015 | 522 | TCH |
| Anonymous hospital No. 4 | 2012-2015 | 324 | SCH |
| Anonymous hospital No. 5 | 2015 | 258 | SCH |
| Anonymous hospital No. 6 | 2012 | 428 | SCH |
| Anonymous hospital No. 7 | 2012-2015 | 310 | SCH |
| Anonymous hospital No. 8 | 2013, 2014, 2015 | 254 | SCH |
| Anonymous hospital No. 9 | 2012-2015 | 315 | SCH |
| Anonymous hospital No. 10 | 2012-2015 | 832 | TCH |
| Anonymous hospital No. 11 | 2012-2015 | 555 | TCH |
| Anonymous hospital No. 12 | 2012-2013, 2015 | 755 | TCH |
| Anonymous hospital No. 13 | 2012-2015 | 503 | SCH |
| Anonymous hospital No. 14 | 2012, 2014 | 314 | SCH |
| Anonymous hospital No. 15 | 2012-2015 | 625 | TCH |
| Anonymous hospital No. 16 | 2012-2015 | 697 | TCH |
| Anonymous hospital No. 17 | 2012-2015 | 1000 | TCH |
| Anonymous hospital No. 18 | 2012 | 444 | SCH |
| Anonymous hospital No. 19 | 2012-2015 | 867 | TCH |
| Anonymous hospital No. 20 | 2012-2015 | 324 | SCH |
| Anonymous hospital No. 21 | 2012-2015 | 349 | SCH |
| Anonymous hospital No. 22 | 2012-2015 | 549 | SCH |
| Anonymous hospital No. 23 | 2014-2015 | 505 | SCH |
| Anonymous hospital No. 24 | 2012-2015 | 600 | TCH |
| Anonymous hospital No. 25 | 2012-2015 | 306 | SCH |
| Anonymous hospital No. 26 | 2012-2015 | 260 | SCH |
| Anonymous hospital No. 27 | 2013-2015 | 411 | SCH |
| Anonymous hospital No. 28 | 2014-2015 | 395 | SCH |
| Anonymous hospital No. 29 | 2012-2015 | 420 | SCH |
| Anonymous hospital No. 30 | 2012-2015 | 232 | SCH |
| Anonymous hospital No. 31 | 2012-2015 | 360 | SCH |
| Anonymous hospital No. 32 | 2015 | 150 | SCH |
| Anonymous hospital No. 33 | 2012-2015 | 672 | TCH |
| Anonymous hospital No. 34 | 2014-2015 | 334 | SCH |
| Anonymous hospital No. 35 | 2012 | 320 | SCH |
| Anonymous hospital No. 36 | 2012-2015 | 307 | SCH |
| Anonymous hospital No. 37 | 2012-2015 | 878 | TCH |
| Anonymous hospital No. 38 | 2012-2015 | 344 | SCH |

| | | | |
|---|---|---|---|
| Anonymous hospital No. 39 | 2012-2015 | 420 | SCH |
| Anonymous hospital No. 40 | 2012-2015 | 360 | SCH |
| Anonymous hospital No. 41 | 2012-2013, 2015 | 210 | SCH |
| Anonymous hospital No. 42 | 2012-2015 | 602 | TCH |
| Anonymous hospital No. 43 | 2012-2015 | 560 | TCH |
| Anonymous hospital No. 44 | 2012-2015 | 863 | TCH |
| Anonymous hospital No. 45 | 2012-2015 | 215 | SCH |
| Anonymous hospital No. 46 | 2012-2015 | 509 | SCH |
| Anonymous hospital No. 47 | 2012-2015 | 480 | SCH |
| Anonymous hospital No. 48 | 2012-2015 | 335 | SCH |
| Anonymous hospital No. 49 | 2012-2015 | 527 | TCH |

* SCH=Secondary-care hospital; TCH=Tertiary-care hospital

**Supplementary File 3. Proportion of AMR BSI**

| Pathogens* | Tertiary-care hospitals (n=15 hospitals) | Secondary-care hospitals (n=34 hospitals) | P value** |
|---|---|---|---|
| Community-origin BSI | | | |
| CRAB | 36.9% (444/1,203) | 25.0% (139/557) | 0.041 |
| CRPA | 18.4% (224/1,218) | 17.1% (140/821) | 0.44 |
| 3GCREC | 37.7% (4,462/11,845) | 31.2% (3,110/9,954) | 0.020 |
| 3GCRKP | 24.4% (957/3,930) | 19.2% (538/2,805) | 0.053 |
| CREC | 0.9% (90/9,871) | 1.6% (136/8,597) | 0.27 |
| CRKP | 2.3% (74/3,265) | 2.7% (62/2,318) | 0.53 |
| MRSA | 11.7% (438/3,733) | 11.2% (291/2,598) | 0.46 |
| Hospital-origin BSI | | | |
| CRAB | 75.5% (2,290/3,035) | 68.3% (697/1,021) | 0.40 |
| CRPA | 36.7% (474/1,292) | 36.7% (203/553) | 0.76 |
| 3GCREC | 53.9% (1,495/2,774) | 53.4% (703/1,313) | 0.25 |
| 3GCRKP | 63.7% (1,500/2,356) | 57.3% (538/939) | 0.14 |
| CREC | 3.0% (66/2,230) | 3.2% (38/1,176) | 0.69 |
| CRKP | 9.2% (196/2,124) | 8.6% (73/849) | 0.84 |
| MRSA | 37.4% (634/1,695) | 28.0% (249/891) | 0.28 |

* Community-origin (Hospital-origin) BSI was defined for patients in the hospital within (longer than) the first two calendar days of admission when the first blood specimen culture positive for a pathogen were taken, with calendar day one equal to the day of admission. The proportion of AMR was defined as new AMR BSI over all patients with new BSIs for the pathogen of interest. Those who were not tested for the antibiotic of interest were not included. **The comparison was made using mixed-effect logistic regression models of patients nested within hospitals.

**Supplementary File 4. Frequency of AMR BSI per 100,000 admissions**

| Pathogens* | Tertiary-care hospitals (n of admissions = 3,134,815) | Secondary-care hospitals (n of admissions = 2,867,762) | Incidence rate ratio (95% confidence interval)** | P value** |
|---|---|---|---|---|
| Community-origin BSI (per 100,000 admissions) | | | | |
| CRAB | 14.2 (n=444) | 4.8 (n=139) | 2.92 (1.87-4.56) | <0.0001 |
| CRPA | 7.1 (n=224) | 4.9 (n=140) | 1.49 (0.88-2.53) | 0.062 |
| 3GCREC | 142.3 (n=4,462) | 108.4 (n=3,110) | 1.33 (1.03-1.73) | 0.070 |
| 3GCRKP | 30.5 (n=957) | 18.8 (n=538) | 1.57 (1.17-2.11) | 0.0017 |
| CREC | 2.9 (n=90) | 4.7 (n=136) | 0.68 (0.39-1.17) | 0.16 |
| CRKP | 2.4 (n=74) | 2.2 (n=62) | 1.10 (0.59-2.03) | 0.77 |
| MRSA | 14.0 (n=438) | 10.1 (n=291) | 1.42 (0.81-2.47) | 0.22 |
| Hospital-origin BSI (per 100,000 admissions) | | | | |
| CRAB | 73.1 (n=2,290) | 24.3 (n=697) | 3.10 (1.90-5.06) | <0.0001 |
| CRPA | 15.1 (n=474) | 7.1 (n=203) | 2.42 (1.36-4.32) | 0.0026 |
| 3GCREC | 47.7 (n=1,495) | 24.5 (n=703) | 1.99 (1.40-2.82) | 0.0001 |
| 3GCRKP | 47.8 (n=1,500) | 18.7 (n=538) | 2.49 (1.70-3.64) | <0.0001 |
| CREC | 2.1 (n=66) | 1.3 (n=38) | 1.66 (0.77-3.58) | 0.20 |
| CRKP | 6.3 (n=196) | 2.5 (n=73) | 2.35 (1.10-5.00) | 0.027 |
| MRSA | 20.2 (n=634) | 8.7 (n=249) | 2.12 (1.30-3.45) | 0.0025 |

* Community-origin (Hospital-origin) BSI was defined for patients in the hospital within (longer than) the first two calendar days of admission when the first blood specimens culture positive for a pathogen were taken, with calendar day one equal to the day of admission. **The comparison was made using mixed-effect Poisson regression models of patients nested within hospitals.

**Supplementary File 5. Frequency of AMR BSI per 100,000 patient-days at risk**

| Pathogens* | Tertiary-care hospitals (n of patient-days at risk = 12,341,585) | Secondary-care hospitals (n of patient-days at risk = 9,988,198) | Incidence rate ratio (95% confidence interval)** | P value** |
|---|---|---|---|---|
| Hospital-origin BSI (per 100,000 patient-days at risk) | | | | |
| CRAB | 18.6 (n=2,290) | 7.0 (n=697) | 2.77 (1.72-4.43) | <0.0001 |
| CRPA | 3.8 (n=474) | 2.0 (n=203) | 2.14 (1.22-3.74) | 0.0073 |
| 3GCREC | 12.1 (n=1,495) | 7.0 (n=703) | 1.79 (1.29-2.50) | 0.0005 |
| 3GCRKP | 12.2 (n=1,500) | 5.4 (n=538) | 2.23 (1.57-3.17) | <0.0001 |
| CREC | 0.5 (n=66) | 0.4 (n=38) | 1.41 (0.65-3.06) | 0.39 |
| CRKP | 1.6 (n=196) | 0.7 (n=73) | 2.10 (1.01-4.35) | 0.045 |
| MRSA | 5.1 (n=634) | 2.5 (n=249) | 1.88 (1.17-3.01) | 0.0091 |

* Hospital-origin BSI was defined for patients in the hospital longer than the first two calendar days of admission when the first blood specimens culture positive for a pathogen were taken, with calendar day one equal to the day of admission. Patients were considered at risk of hospital-origin BSI after they stayed in the hospital for more than two calendar days. **The comparison was made using mixed-effect Poisson regression models of patients nested within hospitals.

**Supplementary File 6. Frequency of AMR BSI per 100,000 tested patients**

| Pathogens* | Tertiary-care hospitals (n of tested patients = 191,399) | Secondary-care hospitals (n of tested patients = 206,572) | Incidence rate ratio (95% confidence interval)** | P value** |
|---|---|---|---|---|
| Community-origin BSI (per 100,000 tested patients) | | | | |
| CRAB | 107.1 (n=150) | 49.4 (n=102) | 2.22 (1.26-3.92) | 0.006 |
| CRPA | 54.3 (n=104) | 47.0 (n=97) | 1.23 (0.57-2.69) | 0.59 |
| 3GCREC | 1191.8 (n=2,281) | 1119.7 (n=2,313) | 0.97 (0.64-1.49) | 0.91 |
| 3GCRKP | 274.3 (n=525) | 179.6 (n=371) | 1.43 (0.99-2.07) | 0.059 |
| CREC | 24.0 (n=46) | 55.2 (n=114) | 0.50 (0.22-1.12) | 0.091 |
| CRKP | 20.9 (n=40) | 23.7 (n=49) | 0.86 (0.37-2.03) | 0.73 |
| MRSA | 133.2 (n=255) | 107.5 (n=222) | 1.25 (0.66-2.39) | 0.48 |
| | Tertiary-care hospitals (n of tested patients = 50,603) | Secondary-care hospitals (n of tested patients = 41,441) | Incidence rate ratio (95% confidence interval)** | P value** |
| Hospital-origin BSI (per 100,000 tested patients) | | | | |
| CRAB | 1,991.3 (n=733) | 1,201.7 (n=498) | 1.89 (1.06-3.37) | 0.032 |
| CRPA | 584.9 (n=296) | 345.1 (n=143) | 1.81 (0.91-3.61) | 0.091 |
| 3GCREC | 1,671.8 (n=846) | 1,158.3 (n=480) | 1.43 (0.97-2.09) | 0.069 |
| 3GCRKP | 1,774.6 (n=898) | 851.8 (n=353) | 2.10 (1.47-3.00) | <0.0001 |
| CREC | 83.0 (n=42) | 74.8 (n=31) | 1.10 (0.52-2.36) | 0.81 |
| CRKP | 252.9 (n=128) | 113.4 (n=47) | 2.32 (1.07-5.03) | 0.033 |
| MRSA | 743.0 (n=376) | 381.3 (n=158) | 1.72 (1.00-2.96) | 0.049 |

* Community-origin (Hospital-origin) BSI was defined for patients in the hospital within (longer than) the first two calendar days of admission when the first blood specimens culture positive for a pathogen were taken, with calendar day one equal to the day of admission. Frequency of AMR BSI per 100,000 tested patients were estimated in 23 SCHs and 7 TCHs of which the microbiology data obtained included the culture result of "no growth". **The comparison was made using mixed-effect Poisson regression models of patients nested within hospitals.

**Supplementary File 7. In-hospital mortality of patients with AMR BSI**

| Pathogens * | Tertiary-care hospitals (n=15 hospitals) | Secondary-care hospitals (n=34 hospitals) | P value ** |
|---|---|---|---|
| Community-origin BSI | | | |
| CRAB | 48.0% (213/444) | 47.5% (66/139) | 0.93 |
| CRPA | 26.8% (60/224) | 30.0% (42/140) | 0.95 |
| 3GCREC *** | 23.8% (1,048/4,399) | 22.5% (681/3,032) | 0.46 |
| 3GCRKP *** | 30.8% (275/894) | 28.3% (140/495) | 0.28 |
| CREC | 28.9% (26/90) | 29.4% (40/136) | 0.93 |
| CRKP | 37.8% (28/74) | 27.4% (17/62) | 0.20 |
| MRSA | 33.3% (146/438) | 31.3% (91/291) | 0.83 |
| Hospital-origin BSI | | | |
| CRAB | 55.1% (1,261/2,290) | 51.9% (362/697) | 0.88 |
| CRPA | 42.6% (202/474) | 40.9% (83/203) | 0.86 |
| 3GCREC *** | 34.6% (496/1,434) | 35.0% (236/675) | 0.88 |
| 3GCRKP *** | 38.1% (497/1,306) | 37.3% (176/472) | 0.74 |
| CREC | 39.4% (26/66) | 42.1% (16/38) | 0.71 |
| CRKP | 49.0% (96/196) | 41.1% (30/73) | 0.38 |
| MRSA | 44.6% (283/634) | 43.8% (109/249) | 0.75 |

* Community-origin (Hospital-origin) BSI was defined for patients in the hospital within (longer than) the first two calendar days of admission when the first blood specimens culture positive for a pathogen were taken, with calendar day one equal to the day of admission. In-hospital mortality (%) of AMR BSI was calculated as the percentage of patients with new AMR BSI who died in the hospitals. Those who were not tested for the antibiotic of interest were not included. **The comparison was made using mixed-effect logistic regression models of patients nested within hospitals. *** For the analysis of mortality, patients with 3GCREC BSI did not include patients with CREC BSI, and patients with 3GCRKP did not include patients with CRKP BSI.

**Supplementary File 8. Mortality rate of AMR BSI per 100,000 admissions**

| Pathogens * | Tertiary-care hospitals (n of admissions = 3,134,815) | Secondary-care hospitals (n of admissions = 2,867,762) | Mortality rate ratio (95% confidence interval) ** | P value ** |
|---|---|---|---|---|
| Community-origin BSI (per 100,000 admissions) | | | | |
| CRAB | 6.8 (n=213) | 2.3 (n=66) | 2.92 (1.65-5.19) | <0.0001 |
| CRPA | 1.9 (n=60) | 1.5 (n=42) | 1.45 (0.70-3.00) | 0.32 |
| 3GCREC *** | 33.4 (n=23) | 23.7 (n=681) | 1.40 (0.97-2.01) | 0.069 |
| 3GCRKP *** | 8.8 (n=275) | 4.9 (n=140) | 1.79 (1.28-2.52) | <0.0001 |
| CREC | 0.8 (n=26) | 1.4 (n=40) | 0.62 (0.33-1.16) | 0.14 |
| CRKP | 0.9 (n=28) | 0.6 (n=17) | 1.54 (0.73-3.27) | 0.25 |
| MRSA | 4.7 (n=146) | 3.2 (n=91) | 1.47 (0.71-3.02) | 0.30 |
| Hospital-origin BSI (per 100,000 admissions) | | | | |
| CRAB | 40.2 (n=1,261) | 12.6 (n=362) | 3.06 (1.84-5.07) | <0.0001 |
| CRPA | 6.4 (n=202) | 2.9 (n=83) | 2.44 (1.24-4.81) | 0.0099 |
| 3GCREC *** | 15.8 (n=496) | 8.2 (n=236) | 1.86 (1.20-2.88) | 0.0056 |
| 3GCRKP *** | 15.9 (n=497) | 6.1 (n=176) | 2.50 (1.60-3.90) | 0.0001 |
| CREC | 0.8 (n=26) | 0.6 (n=16) | 1.48 (0.63-3.48) | 0.37 |
| CRKP | 3.1 (n=96) | 1.0 (n=30) | 2.64 (1.10-6.33) | 0.029 |
| MRSA | 9.0 (n=283) | 3.8 (n=109) | 2.06 (1.19-3.59) | 0.010 |

* Community-origin (Hospital-origin) BSI was defined for patients in the hospital within (longer than) the first two calendar days of admission when the first blood specimens culture positive for a pathogen were taken, with calendar day one equal to the day of admission. **The comparison was made using mixed-effect Poisson regression models of patients nested within hospitals. *** For the analysis of mortality, patients with 3GCREC BSI did not include patients with CREC BSI, and patients with 3GCRKP did not include patients with CRKP BSI.

**Supplementary File 9. Mortality rate of AMR BSI per 100,000 patient-days at risk**

| Pathogens * | Tertiary-care hospitals (n of patient-days at risk = 12,341,585) | Secondary-care hospitals (n of patient-days at risk = 9,988,198) | Mortality rate ratio (95% confidence interval) ** | P value ** |
|---|---|---|---|---|
| Hospital-origin BSI (per 100,000 patient-days at risk) | | | | |
| CRAB | 10.2 (n=1,261) | 3.6 (n=362) | 2.77 (1.71-4.48) | <0.0001 |
| CRPA | 1.6 (n=202) | 0.8 (n=83) | 2.16 (1.13-4.13) | 0.020 |
| 3GCREC *** | 4.0 (n=496) | 2.4 (n=236) | 1.71 (1.14-2.57) | 0.0091 |
| 3GCRKP *** | 4.0 (n=497) | 1.8 (n=176) | 2.24 (1.49-3.38) | <0.0001 |
| CREC | 0.2 (n=26) | 0.2 (n=16) | 1.28 (0.56-2.99) | 0.56 |
| CRKP | 0.8 (n=96) | 0.3 (n=30) | 2.38 (1.03-5.48) | 0.042 |
| MRSA | 2.3 (n=283) | 1.1 (n=109) | 1.84 (1.08-3.11) | 0.023 |

* Hospital-origin BSI was defined for patients in the hospital longer than the first two calendar days of admission when the first blood specimens culture positive for a pathogen were taken, with calendar day one equal to the day of admission. Patients were considered at risk of hospital-origin BSI after they stayed in the hospital for more than two calendar days. **The comparison was made using mixed-effect Poisson regression models of patients nested within hospitals. *** For the analysis of mortality, patients with 3GCREC BSI did not include patients with CREC BSI, and patients with 3GCRKP did not include patients with CRKP BSI.

**Supplementary File 10. Total number and mortality of patients with notifiable bacterial diseases**

| Pathogens* | Total number of patients | Total number of in-hospital deaths | In-hospital mortality (%) |
|---|---|---|---|
| *Burkholderia pseudomallei* | 6,164 | 1,524 | 24.7% |
| *Brucella* spp. | 4 | 0 | 0% |
| *Corynebacterium diphtheriae* | 40 | 9 | 22.5% |
| *Neisseria gonorrhoeae* | 60 | 0 | 0% |
| *Neisseria meningitidis* | 13 | 3 | 23.1% |
| Non-typhoidal *Salmonella* spp. | 11,264 | 1,005 | 8.9% |
| *Salmonella enterica* serovar Paratyphi | 30 | 4 | 13.3% |
| *Salmonella enterica* serovar Typhi | 115 | 4 | 3.5% |
| *Shigella* spp. | 361 | 7 | 1.9% |
| *Streptococcus suis* | 710 | 85 | 12.0% |
| *Vibrio* spp, | 2,143 | 172 | 8.0% |

* Notifiable bacterial diseases were defined as culture positive for pathogens of interest in any type of clinical specimens including blood, cerebrospinal fluid, respiratory tract specimens, urine, genital swab, stool and other or unknown sample types. The AMASS application merged the microbiology data file and hospital admission data file. The merged dataset was then de-duplicated so that only the first isolate per patient per specimen per reporting period was included in the analysis. In-hospital mortality (%) was calculated as the percentage of patients with new AMR BSI who died in the hospitals.
